# The Utility of a Bayesian Predictive Model to Forecast Neuroinvasive West Nile Virus Disease in the United States, 2022

**DOI:** 10.1101/2022.11.02.22281839

**Authors:** Maggie S. J. McCarter, Stella Self, Kyndall C. Dye-Braumuller, Christopher Lee, Huixuan Li, Melissa S. Nolan

**Affiliations:** Department of Epidemiology and Biostatistics, University of South Carolina; Department of Computer Science and Engineering, University of South Carolina

## Abstract

Arboviruses (arthropod-borne-viruses) are an emerging global health threat that are rapidly spreading as climate change, international business transport, and landscape fragmentation impact local ecologies. Since its initial detection in 1999, West Nile virus (WNV) has shifted from being a novel to an established arbovirus in the United States. Subsequently, more than 25,000 cases of West Nile Neuro-invasive Disease (WNND) have been diagnosed, cementing WNV as an arbovirus of public health importance. Given its novelty in the United States, high-risk ecologies are largely underdefined making targeted population-level public health interventions challenging. Using the Centers for Disease Control and Prevention ArboNET WNV data from 2000 – 2021, this study aimed to predict WNND human cases at the county level for the contiguous US states using a spatio-temporal Bayesian negative binomial regression model. The model includes environmental, climatic, and demographic factors, as well as the distribution of host species. An integrated nested LaPlace approximation (INLA) approach was used to fit our model. To assess model prediction accuracy, annual counts were withheld, forecasted, and compared to observed values. The validated models were then fit to the entire dataset for 2022 predictions. This proof-of-concept mathematical, geospatial modelling approach has proven utility for national health agencies seeking to allocate funding and other resources for local vector control agencies tackling WNV and other notifiable arboviral agents.

## 1. Introduction

West Nile virus (WNV) is an arthropod-borne virus (arbovirus) in the family Flaviviridae (genus *Flavivirus*) first isolated in the West Nile district of Uganda in 1937 (Smithburn et al., 1946). Since its introduction into the New World in 1999, it has become widely distributed in North and Central America and is most likely established in South America (Artsob et al., 2009; Marcondes et al., 2017). It is thought that WNV is the most widely distributed arbovirus globally (Reisen, 2013). The primary mosquito vectors of WNV are *Culex* spp., specifically those in the *Culex pipiens* L. complex, *Culex tarsalis* Coquillett, *Culex restuans* Theobald, *Culex nigripalpus* Theobald, and the *Culex univittatus* Theobald complex (Foster & Walker, 2002; Sardelis et al., 2001). WNV is maintained in the environment through an enzootic cycle between these mosquito vectors and Passiformes birds as vertebrate reservoirs and amplifying hosts during epidemics. The virus can be spread to additional vertebrates such as humans when a bridge vector mosquito species feeds on a mammal; however, humans do not produce high enough viremia to continue the spread of the virus into additional mosquitoes with subsequent bites.

Clinical symptoms of disease typically appear in approximately 20% of those infected; these include fever, headache, body aches, joint pains, vomiting, diarrhea, or rash (*Symptoms, Diagnosis, & Treatment* | *West Nile Virus* | *CDC*, 2021). Severe illness affecting the central nervous system with encephalitis and meningitis can occur in <1% of cases, called West Nile neuroinvasive disease (WNND) (Beckham & Tyler, 2015). In the continental United States (US), WNV disease is the leading cause of mosquito-borne illness (*West Nile Virus* | *West Nile Virus* | *CDC*, 2022). Cumulative estimates of its impact have suggested WNV has caused more than 25,000 neuroinvasive disease cases and 7 million infections since its introduction into the US (Ronca et al., 2019). In addition to its public health impacts, WNV has also produced a heavy economic burden, where the cost of acute clinical care and subsequent long-term costs associated with infection is estimated at $56 million annually (Barrett, 2014; Ronca et al., 2021).

With these estimated case and medical cost burdens, the US must stay vigilant and active in mosquito and mosquito-borne disease surveillance. The National Association of County and City Health Officials (NACCHO) published a national Vector Control Assessment in 2020, finding that less than a quarter of mosquito and vector control programs in the US are “fully capable” regarding mosquito surveillance and control capacity (*New Report Reveals State of Local Vector Control Capacity in the U*.*S. - NACCHO*, n.d.). Additional regional and state vector control capacity surveys have reported similar results: the majority of mosquito and vector control agencies or programs are not prepared to handle major vector-borne disease outbreaks or are not proactive in various surveillance capabilities, leaving the US vulnerable (Dye-Braumuller et al., 2022; Moise et al., 2020; Peper et al., 2022). This vulnerability, in combination with anthropogenic climate change, urbanization, and international import and export, has set the stage for increased arboviral transmission in the US (Keyel et al., 2021; Wimberly et al., 2020). Clearly, empirical knowledge of disease cases and vector species infection is not enough to prevent disease. Instead, vector-borne disease forecast modeling, relying on input from a multitude of disciplines including epidemiology, entomology, ecology, and biology to predict risk can be used to prevent continued spread and additional outbreaks.

Mathematical and computational modeling for epidemiology has significantly advanced in recent years and is commonly used for its insight into spatio-temporal transmission dynamics, synthesis of multi-disciplinary inputs, and overall reduction in cost compared to traditional surveillance (Siettos & Russo, 2013). Given the zoonotic nature of many vector-borne diseases, these modeling techniques are particularly useful when traditional disease surveillance cannot reach all potential animal reservoirs or hosts. Specifically for WNV, multiple methods of forecasting have been performed. Bayesian models fit using Markov chain Monte Carlo (MCMC) methods have been heavily utilized for decades in disease prediction, and have shown to be effective for the prediction of WNV and corresponding associations(Foppa et al., 2011; O’Neill et al., 2000; Temple et al., 2022). However, MCMC approaches are often computationally expensive. The Integrated Nested Laplace Approximation (INLA) technique is a streamlined alternative to the traditional MCMC(Rue et al., 2009). Contrary to the MCMC approach of estimating the joint posterior distribution of the parameters, the INLA model uses marginal inference on individual posteriors. This greatly reduces computation time while remaining precise in estimation.

In May 2022, the CDC released a call for predictions of WNND for the year 2022 to predict disease for more efficient allocation of resources and preparedness for this vector-borne disease. This paper describes one prediction model developed for this challenge to ultimately predict cases of WNND at the county level for the contiguous US states using a spatio-temporal Bayesian negative binomial regression, fit using an INLA approach. This is one of the first studies to predict human mosquito-borne disease for the contiguous US at the county level.

## 2. Methods

### 2.1 WNND Data

Data on WNND were obtained from the Centers for Disease Control and Prevention’s ArboNET, a national arboviral surveillance database maintaining data on arboviral infections in humans, veterinary disease, and vector and host prevalence and health. ArboNET human cases are collected from clinicians who diagnose patients with an arboviral disease, and include both neuroinvasive and non-neuroinvasive WNV cases (Centers for Disease Control and Prevention, 2022). Only neuroinvasive cases were included in this study.

### 2.2 Covariates

Covariates were selected through a thorough literature search. We obtained the population aged 65 years of age and older from each county from the US Census Bureau. Avian host data was collected from the eBird database, a global citizen-science database dedicated to facilitating understanding of avian patterns (Sullivan et al., 2009). Data was obtained for all sightings of crows, jays, and sparrows in the contiguous US from January 1, 2000 to December 31, 2021, and linear interpolation was used to estimate avian population for 2022. Available data included the latitude-longitude location of the sighting and the number of birds sighted. To account for the fact that some locations are visited by bird watchers more often than others, we averaged the number of birds sighted per location per observer. Outliers with extreme values were removed and kriging was used to estimate the mean number of birds sighted at each county centroid. Land cover data was obtained from the National Land Cover Database (NLCD), a national dataset that characterizes landcover status and changes from 2001, 2006, 2011, and 2019. Linear interpolation was performed to assess land cover information for the years for which NLCD was not available. For each county, the proportion of that county’s land mass which fell into each of 14 NLCD categories was computed. Our analysis accounted for el Niño, an unusual warmth pattern in international climate, presence, and absence across the timeline. El Niño years were collected from the National Oceanic and Atmospheric Administration’s Climate Prediction Center. We created a binary “el Niño” variable as a 0 or 1 value for el Niño year and included the variable in our final dataset. County level data on the presence/absence *Culex quinquifasciatus* mosquito presence were obtained from data provided by Wang et al., and was assumed to be static over all study years. (Wang et al., 2014). Finally, as WNV cases tend to exhibit a three-year cycle of increase and decrease, indicator variables for cycle-year (cycle year 1, cycle year 2, or cycle year 3 (reference level) were included in the model.

### 2.3 The Model

To avoid over-dispersion, we utilized a Bayesian negative binomial regression model. Our model can be described via the following equations:

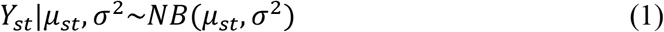

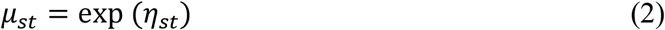

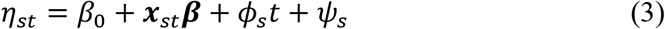

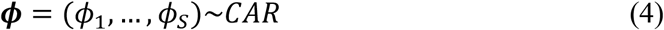

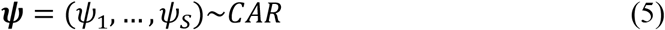

The number of cases in county s and year t are signified by *Y*_*st*_, and *Y*~*NB*(*μ,σ*^2^) indicates that the random variable *Y* follows a negative binomial distribution with mean μ and overdispersion parameter *σ*^2^. The mean *μ*_*st*_ is related to a linear *η*_*st*_ which is a function of covariates; σ^2^ is a non-negative overdispersion parameter. Four pieces compose *η*_*st*_: the shared intercept term *β*_0_, covariate factors ***x***_*st*_, a spatially varying temporal trend parameter *ϕ*_*t*_, and a spatially varying intercept *ψ*_*s*_. Because the effect of time varied across regions, we used random effects to allow the coefficients for year to be county-specific and included a spatial random effect intercept. These random effects (***ϕ*** and ***ψ***) are assumed to follow conditional auto-regressive (CAR) priors to ensure that random effects from counties that are close in space have similar values, as counties that are adjacent to each other often have similar incidence rate and similar trends of increase/decrease. For more on CAR models, see Banerjee et. Al 2003 or Besag 1974 (Banerjee et al., 2003; Besag, 1974). All other model parameters followed the default prior specification implemented by the R INLA package (Rue et al., 2009).

### 2.4 Model Selection

Once all data on covariates were collected, tests for multicollinearity were performed. Variables with the highest variance inflation factor (VIF) were eliminated until every VIF was below 5. This resulted in removing the variable proportion of land mass in cultivated crop land cover. We then fit the model using all the remaining variables. Next, variables that were not found to be statistically important were removed individually until all variables were statistically important. A variable was deemed statistically important if its 95% credible interval did not contain 0. We then added the removed variables back in individually, calculated the mean and median square prediction error, and kept those that improved the mean square prediction error.

Predictors chosen for the final model included: population of those above 65, proportion of land mass in open water, developed medium intensity, developed high intensity, barren land, deciduous forest, evergreen forest, mixed forest, shrub scrub, herbaceous, and woody wetlands, sparrow, jay, and crow host populations, a binary variable for el Niño year, year, and cycle variables. We considered having a population offset term; however, including the term resulted in greater error in prediction performance. Because of this, we opted to not include an offset term in our final model. An integrated nested LaPlace approximation (INLA) model in R software was used to fit our model (Rue et al., 2009). This provided a faster way to fit our model than standard Markov chain Monte Carlo methods. Additionally, all map figures were created through ArcGIS Pro 2.8.7. The predictive power of our final model was assessed by predicting counts for each year between 2010 and 2021 using data up to but not including that year and comparing the predicted counts to the observed counts for each year.

## 3. Results

Model selection was performed using holdout data for the year 2021. After model selection, our final model was able to predict 2021 cases with a median square prediction error of 0.006 cases^2^. The posterior mean estimates, standard deviations, and 95% credible interval bounds for the fixed effects for our final model can be found in Table 1.

**Table 1:** Estimation with Fixed Effects

| Variable | Estimate | Standard Deviation | Lower Bound of 95% Credible Interval | Upper Bound of 95% Credible Interval |
| --- | --- | --- | --- | --- |
| Intercept | 3.44E+01 | ±4.66E+00 | 2.53E+01 | 4.35E+01 |
| Population 65+* | 1.02E-05 | ±8.05E-07 | 8.59E-06 | 1.17E-05 |
| Open Water* | -1.43E-02 | ±3.87E-03 | -2.19E-02 | -6.75E-03 |
| Developed Medium Intensity* | 1.54E-01 | ±1.75E-02 | 1.19E-01 | 1.88E-01 |
| Developed High Intensity* | -6.81E-02 | ±2.31E-02 | -1.13E-01 | -2.28E-02 |
| Barren Land | 1.92E-02 | ±2.16E-02 | -2.33E-02 | 6.15E-02 |
| Deciduous Forest* | -3.60E-02 | ±3.39E-03 | -4.27E-02 | -2.94E-02 |
| Evergreen Forest* | -2.57E-02 | ±3.70E-03 | -3.29E-02 | -1.85E-02 |
| Mixed Forest* | -2.88E-02 | ±7.75E-03 | -4.40E-02 | -1.36E-02 |
| Shrub Scrub* | -8.29E-03 | ±3.05E-03 | -1.43E-02 | -2.35E-03 |
| Herbaceous | 2.21E-04 | ±3.06E-03 | -5.81E-03 | 6.20E-03 |
| Woody Wetlands* | -1.27E-02 | ±5.73E-03 | -2.40E-02 | -1.55E-03 |
| el Niño * | 1.42E-01 | ±2.81E-02 | 8.66E-02 | 1.97E-01 |
| Sparrow | 2.76E-03 | ±2.14E-03 | -1.44E-03 | 6.97E-03 |
| Crow | 1.10E-03 | ±8.55E-04 | -5.73E-04 | 2.78E-03 |
| Jay | -3.38E-06 | ±7.27E-04 | -1.34E-03 | 1.51E-03 |
| Cycle Variable 1* | -6.69E-01 | ±3.19E-02 | -7.32E-01 | -6.06E-01 |
| Cycle Variable 2* | -4.94E-01 | ±3.03E-02 | -5.53E-01 | -4.35E-01 |
| Year* | -1.80E-02 | ±2.32E-03 | -2.25E-02 | -1.34E-02 |
| *Found Statistically Important |  |  |  |  |

We then compared the fitted values with the observed counts for the specific years. A handful of counties had a mean square prediction error greater than 1,000 counts^2^. This represented a skewed error distribution and therefore we reported median square prediction error.

Model median square prediction error for years 2010 – 2021, and counties with over 100 count difference between observed and expected per year, can be seen in Table 2. For all years, the mean was noticeably larger than the median, which shows evidence of a skewed distribution. The prediction model performed well for most of the counties, with only 32 (1%) of counties with an absolute error of two counts or higher for the year 2021. In 32 counties with errors of two and above, the model tended to noticeably overestimate the number of cases in the those with very high populations.

**Table 2:** Model Median Square Prediction Error and Counties with Highest Error, by Year

| Year | Median Square<br>Prediction Error<br>(Cases<sup>2</sup>) | Counties with Error > 100 Counts |
| --- | --- | --- |
| 2010 | 0.599 | - |
| 2011 | 0.660 | - |
| 2012 | 0.002 | Los Angeles, CA; Dallas, TX |
| 2013 | 0.001 | Los Angeles, CA; Maricopa, AZ |
| 2014 | 0.005 | Los Angeles, CA; Maricopa, AZ; Harris, TX; Cook, IL; Orange, CA |
| 2015 | 0.009 | Los Angeles, CA; Maricopa, AZ; Harris, TX; Cook, IL |
| 2016 | 0.002 | Los Angeles, CA; Maricopa, AZ; Harris, TX; Cook, IL |
| 2017 | 0.004 | Los Angeles, CA; Maricopa, AZ; Harris, TX; Cook, IL |
| 2018 | 0.010 | Los Angeles, CA; Maricopa, AZ; Harris, TX; Cook, IL |
| 2019 | 0.002 | Los Angeles, CA; Maricopa, AZ; Harris, TX; Cook, IL |
| 2020 | 0.003 | Los Angeles, CA; Maricopa, AZ; Harris, TX; Cook, IL |
| 2021 | 0.006 | Los Angeles, CA; Maricopa, AZ; Harris, TX; Cook, IL |

As can be seen in Figure 2, which represents the prediction error as the difference between observed incidence and predicted incidence, our model did well in most counties. However, our model can be improved in counties with large populations. Finally, after model prediction power was assessed with the holdout data, the model was then fit to the entire dataset to make predictions for 2022. The distribution of this prediction can be seen in Figure 3. The lower and upper bounds of the 95% prediction credible interval can be seen in Figures 4 and 5.

**Figure 1:**
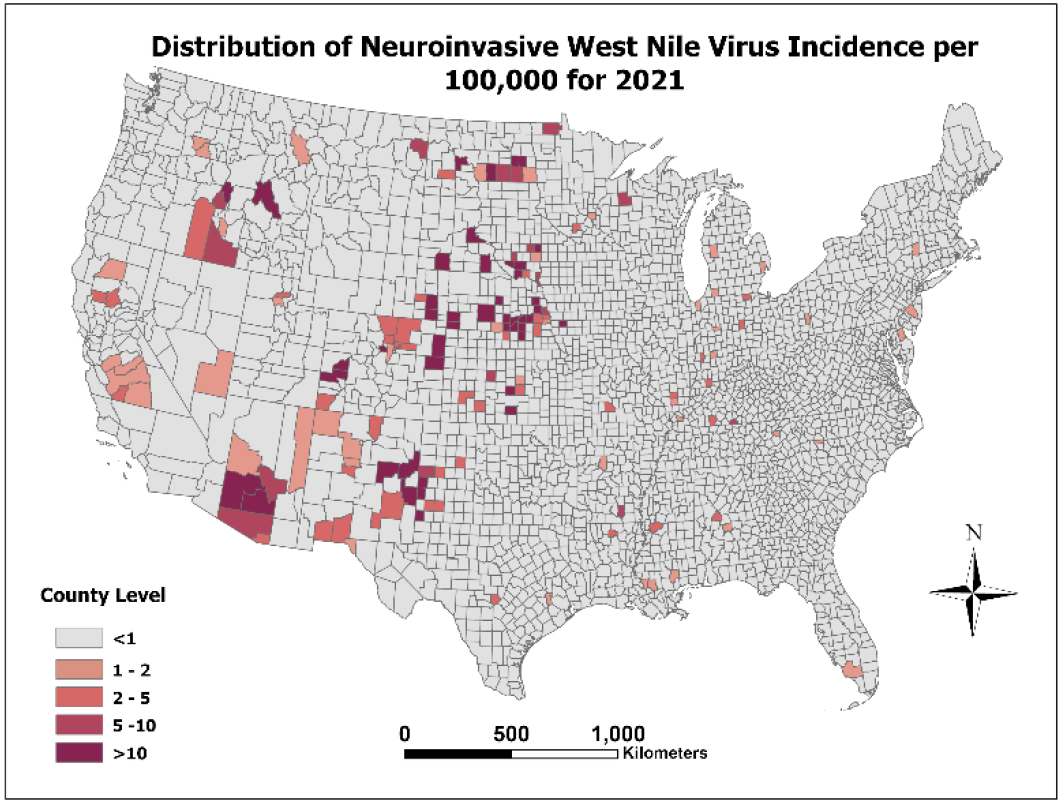
WNND Incidence per 100,000 for the Year 2021

**Figure 2:**
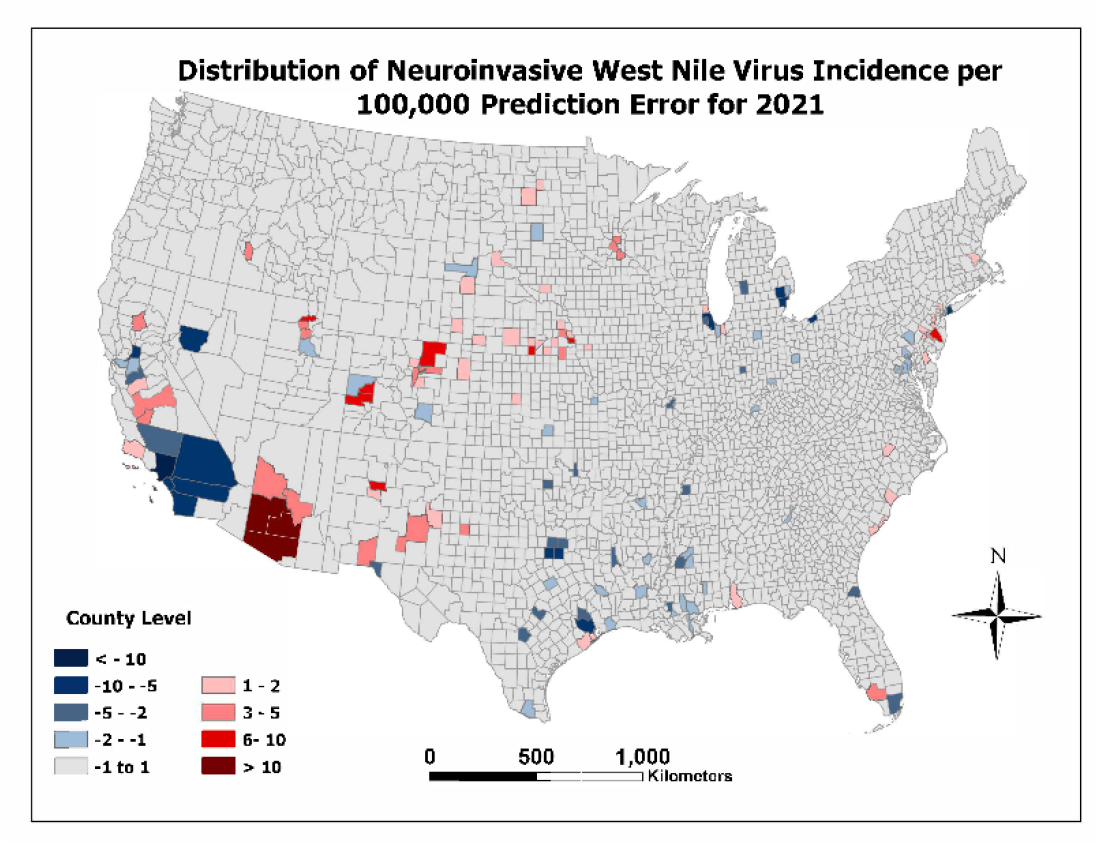
WNND Incidence per 100,000 Prediction Error for the Year 2021

**Figure 3:**
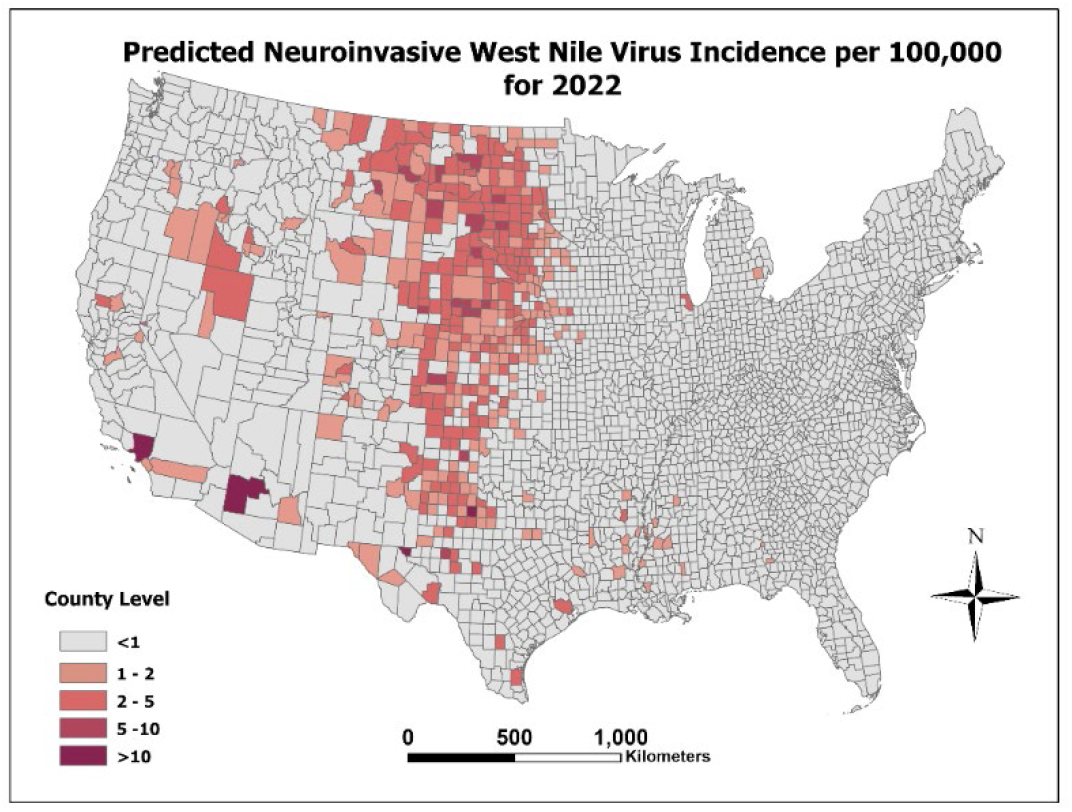
2022 Neuroinvasive West Nile Disease Incidence per 100,000

**Figure 4:**
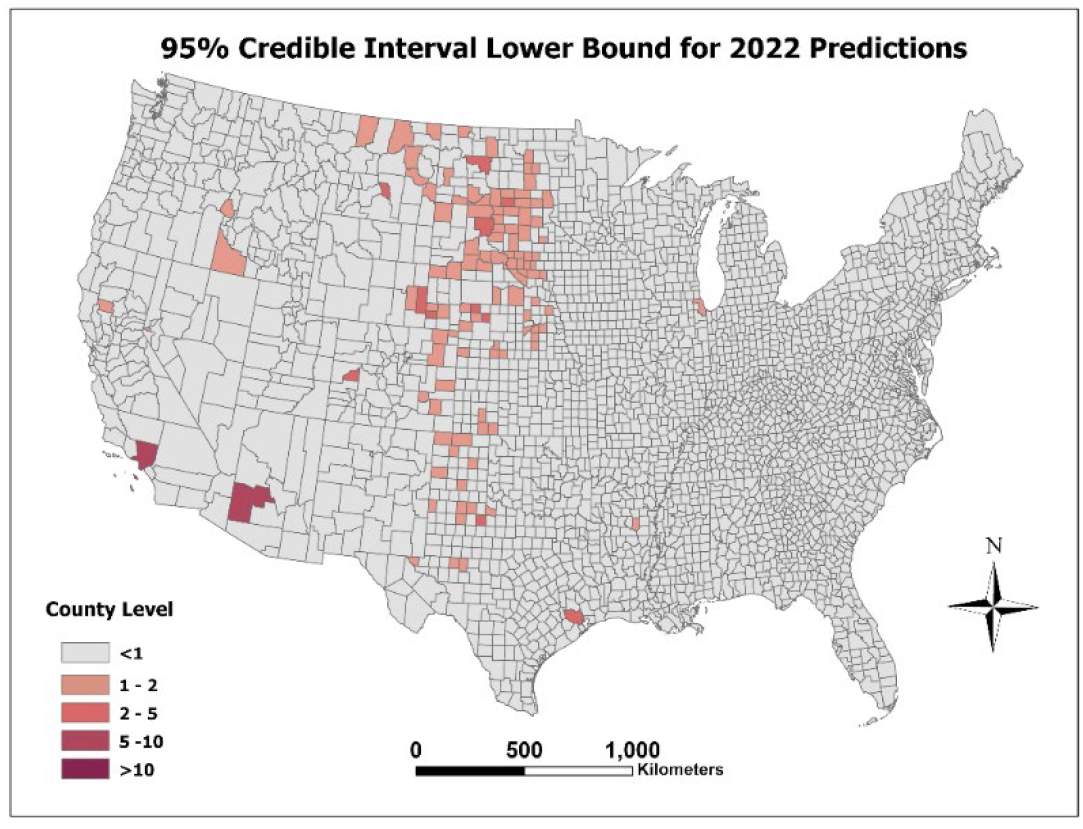
95% Credible Interval Lower Bound for 2022 Predictions

**Figure 5:**
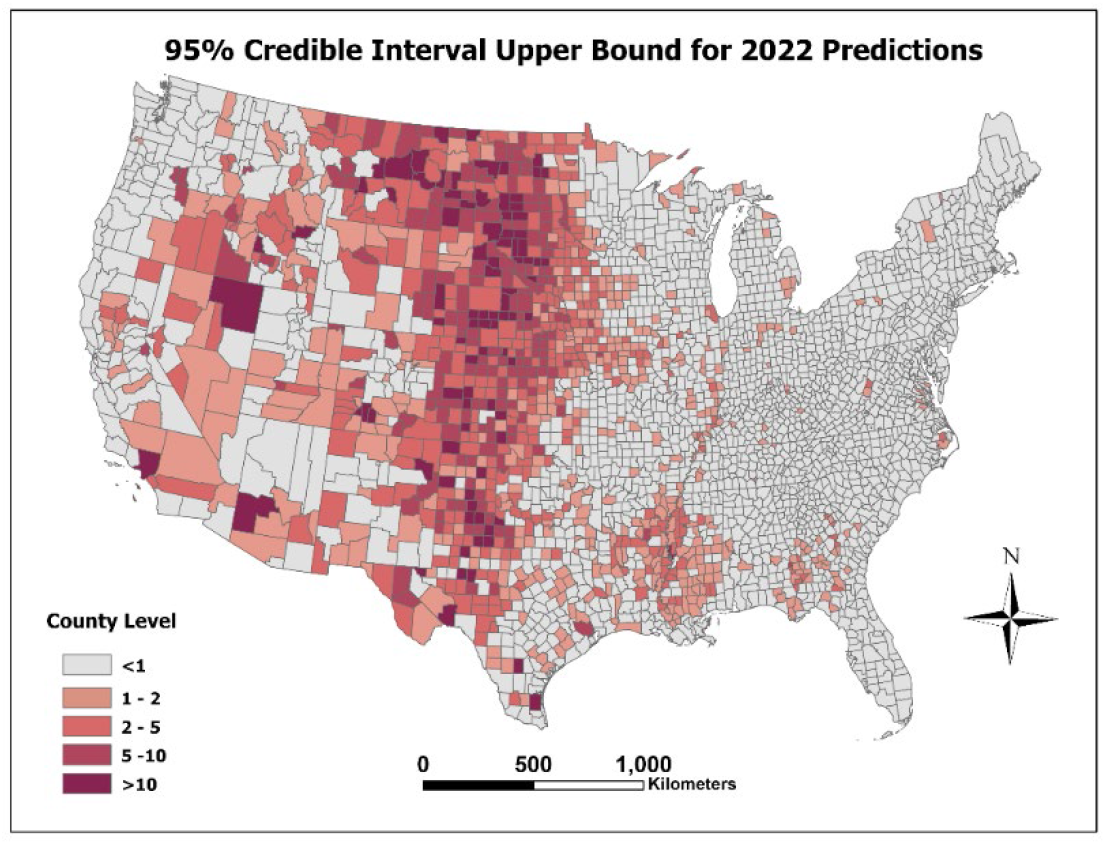
95% Credible Interval Upper Bound for 2022 Predictions

As shown in Figures 3 through 5, the majority of predicted incidence is clustered around the central US. The counties with the highest predicted neuroinvasive WNND incidence were Maricopa, AZ; Los Angeles, CA; Cook, IL; and Harris County, TX, all of which are the top four highest population counties in the United States. These were also the counties that our model overestimated for the 2021 holdout data.

## 4. Discussion and Conclusion

Our 2022 prediction results estimated WNND incidence clustered around the central US. This coincides with most yearly WNND patterns for the last decade, though the years 2016 and 2017 saw a more geographically dispersed WNND incidence, and 2021 showed a more sparce dispersion of WNND incidence. Though the 2022 incidence rates cover a large, concentrated area, the model predicted small rates for most counties. This is contrary to the observed incidence of 2021 where incidence was more dispersed than in 2022, incidence per each county was higher. As evidenced in the supplementary video, WNND incidence ebbs and flows on a 3-year cycle, with incidence peaking in the second year. The year 2021 was a cycle peak-year, and thus 2022 is expected to have a diminishing incidence comparatively. Our predictions confirm this pattern, as WNND incidence in 2022 for most counties is predicted to be less than that of 2021.

The results of this 2022 predictive model can guide government organizations to target high risk areas for prevention efforts and allocation of resources for education programs, mosquito surveillance, and mosquito-borne disease surveillance. Open water, forest landcover, and woody wetland variables were found to have a negative association with WNND counts. This could be an effect of rural communities having less access to healthcare and thus, fewer diagnoses/reports of WNND. Woody wetlands typically support ephemeral breeding or marsh mosquitoes like *Psorophora* and *Aedes* spp., thus there may not be as many *Culex* spp. appropriate breeding habitats here. Open water sources such as ponds or lakes are generally too large to be suitable *Culex* breeding habitats, as they support larger fish and many natural mosquito predators (Stratman, 2008). Developed medium intensity landcover, often associated with suburban areas, was found to have a positive association with WNND counts. This could be due to an environment where mosquitos can easily breed in lawns and standing water, while having a multitude of humans to infect. In contrast, developed high intensity, which can be surmised as concentrated urban areas, were found to have a negative association with WNND counts.

Although there are large populations in cities, there is limited still water or green space for mosquitos to inhabit and breed, making the transmission of WNV less likely. Population 65 years of age and above was found to have a positive relationship with WNND counts. This is to be expected, as WNND has typically been found to affect older populations (McDonald, 2021). El Niño year was found to be positively associated with WNND counts. This coincides with other research linking el Niño to increased infectious disease rates, including WNV (Anyamba et al., 2019). Although the various avian host species prevalence was not found to be statistically important, they greatly improved model mean square prediction error and therefore were kept in the model.

Multiple techniques have been successfully utilized in arboviral disease forecast modelling, including machine learning (Edussuriya et al., 2021), neural networks (Akhtar et al., 2019), and Bayesian models (Myer & Johnston, 2019; Temple et al., 2022). Though neural network and machine learning methods offer accurate predictions, Bayesian modelling adds a probability distribution to these frequency distributions, allowing for seamless inference on predicted counts and other model parameters. This manuscript, utilizing Bayesian inference, is one of the first studies to predict human mosquito borne disease for the entire US at the county level, and introduces concepts that have application for future studies. Because our model showed accurate predictions of WNND, future studies could explore utilizing a similar model to predict non-neuroinvasive WNV incidence in addition to neuroinvasive, or other arboviruses such as dengue virus. Avian vector data has great potential to predict other diseases such as Crimean-Congo hemorrhagic fever virus and tick-borne encephalitis virus. Our model included a yearly-fluctuating “el Niño” variable, but future studies could benefit by developing a more dynamic model over the course of the year including weather data and additional seasonal patterns. Though our model performed well in predicting WNND cases in the contiguous US, it is not without limitations. Diagnoses and reporting of Arboviruses are often incomplete or underreported, therefore incidence of WNND could be underestimated for this study.

Additionally, avian host data was citizen collected and therefore was not standardized across locations in the United States. This required us to smooth the data using kriging. Another limitation was our lack of mosquito vector data at the county level. This was difficult to find, and future studies and models would benefit from obtaining such data. Finally, though accurate in most counties, our model overpredicted counts of WNND in counties with high populations. It is possible that the relationship between population over 65 and WNND cases is non-linear; if such is the case, then the positively estimated linear effect of population over 65 may cause the number of WNND to be drastically overestimated in the counties with very large populations over 65. As these are the same counties with the largest populations overall, this would explain our model’s poor performance in these counties. However, we attempted to address this by including quadratic, cubic, and logarithmic population over 65 terms in the model, but this did not improve model fit. Another potential alternative to fix the over-prediction in high population counties is using a mixture distribution model.

In closing, this study sought to predict cases of neuroinvasive WNV at the county level for the contiguous US states using a spatio-temporal Bayesian negative binomial regression. An integrated nested Laplace approximation approach was utilized to fit the model, saving extensive computational cost. After variable selection, the model showed accurate prediction of historical WNND cases in most counties, though the model can be improved on counties with very large populations. These findings have implications for future stakeholder decisions and interventions regarding targeting areas for WNV prevention.

## Data Availability

All data produced in the present work are available upon reasonable request to the authors.

## Appendix/Supplemental Information

This work was financially supported by the Big Data Health Science Center at the University of South Carolina, and the United States Centers for Disease Control and Prevention (Cooperative Agreement Number U01CK000662). The contents of this publication are solely the responsibility of the authors and do not necessarily represent the official views of the Centers for Disease Control and Prevention or the Department of Health and Human Services. Data was sourced by the CDC’s ArboNET National Arboviral Surveillance System. We would like to acknowledge Dr. Karen Holcomb and Dr. Michael Johansson of the United States Centers for Disease Control and Prevention for their contributions and for their leadership during the 2022 WNND prediction challenge.

## Notes

### Competing Interest Statement

The authors have declared no competing interest.

### Author Declarations

This study used only publicly available West Nile Virus counts from CDC's Arbonet.

